# Dynamics of SARS-CoV-2 exposure in Malawian blood donors: a retrospective seroprevalence analysis between January 2020 and February 2021

**DOI:** 10.1101/2021.08.18.21262207

**Authors:** J Mandolo, J Msefula, MYR Henrion, C Brown, B Moyo, A Samon, T Moyo-Gwete, Z Makhado, F Ayres, T Motlou, N Mzindle, N Kalata, AS Muula, G Kwatra, N Msamala, A Likaka, T Mfune, PL Moore, B Mbaya, N French, RS Heyderman, TD Swarthout, KC Jambo

## Abstract

**Background:** As at end of July 2021, the COVID-19 pandemic has been less severe in sub-Saharan Africa than elsewhere. In Malawi, there have been two subsequent epidemic waves. We therefore aimed to describe the dynamics of SARS-CoV-2 exposure in Malawi.

**Methods:** We measured the seroprevalence of anti-SARS-CoV-2 antibodies among randomly selected blood donor sera in Malawi from January 2020 to February 2021. In a subset, we also assesed *in vitro* neutralisation against the original variant (D614G WT) and the Beta variant.

**Findings:** A total of 3586 samples were selected from the blood donor database, of which 2685 (74.9%) were male and 3132 (87.3%) were aged 20-49 years. Of the total, 469 (13.1%) were seropositive. Seropositivity was highest in October 2020 (15.7%) and February 2021 (49.7%) reflecting the two epidemic waves. Unlike the first wave, both urban and rural areas had high seropositivity by February 2021, Balaka (rural, 37.5%), Blantyre (urban, 54.8%), Lilongwe (urban, 54.5%) and Mzuzu (urban, 57.5%). First wave sera showed potent *in vitro* neutralisation activity against the original variant (78%[7/9]) but not the Beta variant (22% [2/9]). Second wave sera potently neutralised the Beta variant (73% [8/11]).

**Interpretation:** The findings confirm extensive SARS-CoV-2 exposure in Malawi over two epidemic waves with likely poor cross-protection to reinfection from the first on the second wave. Since prior exposure augments COVID-19 vaccine immunity, prioritising administration of the first dose in high SARS-CoV-2 exposure settings could maximise the benefit of the limited available vaccines in Malawi and the region.

**Research in context:** *Evidence before this study:* We searched PubMed on August 16, 2021, with no language restrictions, for titles and abstracts published between Jan 1, 2020, and August 16, 2021, using the search terms: “SARS-CoV-2 seroprevalence in Africa”[Title/Abstract]) OR “SARS-CoV-2 seroprevalence in blood donors” [Title/Abstract] OR “SARS-CoV-2 seroprevalence in Malawi”, and found 15 records. There are limited SARS-CoV-2 seroprevalence studies in sub Saharan Africa, however the few that are available report high seroprevalence than can be deduced from the respective national reported COVID-19 cases and deaths. Only two published SARS-CoV-2 serosurveys were done on blood donors, from Kenya and Madagascar. Blood donor serosurveys have been recommended by the WHO as an important tool for assessing the spread of SARS-CoV-2 and estimating the burden of COVID-19 pandemic.

*Added value of this study:* Unlike previous SARS-CoV-2 blood donor serosurveys in African populations that were conducted for a maximum period of 9 months, our study covers a full year from January 2020 to February 2021, capturing potential introduction of SARS-CoV-2 into Malawi as well as the two epidemic waves. This study provides evidence against the speculation that SARS-CoV-2 had been circulating more widely in sub-Saharan Africa before the first detected cases. It also provides supporting evidence suggesting that the Beta variant was the likely driver of the second wave that resulted in high SARS-CoV-2 seropositivity in January to February 2021 in Malawi.

*Implications of all the available evidence:* Our results show extensive community transmission of SARS-CoV-2 in Malawi as reflected in the blood donors serosurvey, with almost half the sample population being seropositive for anti-SARS-CoV-2 antibodies by February 2021. This has implications for COVID-19 vaccination policy in sub-Saharan Africa (SSA), where there are limited available vaccine doses. Considering that prior exposure to SARS-CoV-2 augments COVID-19 vaccine immunity, strategies to maximise administration of the first vaccine dose, while waiting for more vaccines to become available, could maximise the benefits of the limited available vaccines in high SARS-CoV-2 exposure settings in SSA such as Malawi.

## Introduction

Severe acute respiratory syndrome coronavirus 2 (SARS-CoV-2) emerged in late 2019 in Hubei province, China as a cause of coronavirus disease 2019 (COVID-19) ^1,2^. SARS-CoV-2 infection presents with a range of disease manifestations, from asymptomatic infection to severe acute respiratory distress and death. SARS-CoV-2 has spread globally and the World Health Organisation (WHO) declared COVID-19 a global pandemic on 11^th^ March 2020. As of 12th August 2021, more than 205 million people globally have been infected with more than 4.32 million deaths ^3^. The pandemic has placed substantial pressures on health systems delivering care for patients with COVID-19 and caused disruption of non-COVID-19 health-care provision, in addition to negative effects on the global economy ^4^.

The potential risk from SARS-CoV-2 to Africa was identified early in the global pandemic ^5^. As the epicentre of transmission moved from East Asia to West Asia, Europe and then to North America, there was speculation as to the likely impact of the pandemic on the African continent with its younger populations, high rates of infectious diseases including HIV, poverty and undernutrition, as well as a fragile health care infrastructure ^5,6^. Evidence has shown that the epidemiology of the COVID-19 pandemic in sub-Saharan Africa has been different from the Americas, China and Europe ^3^. Available seroprevalence data indicate that SARS-CoV-2 has circulated more widely in African populations than can be deduced from the number of reported confirmed cases, hospitalisations and deaths ^7-9^. This has led to speculations that SARS-CoV-2 could have circulated longer in sub-Saharan Africa than the first confirmed cases, but current evidence is limited. Others have speculated that a high prevalence of circulating seasonal coronaviruses could have induced some cross-reactive protective immunity ^10,11^.

SARS-CoV-2 Spike and Nucleocapsid specific antibody responses in blood are detectable in individuals between 5 to 21 days post symptom onset ^12^. Subsequently, anti-SARS-CoV-2 IgM antibody levels drop sharply within the initial 14 days followed by a gradual decline in anti-SARS-CoV-2 IgG antibodies within the next 6 months ^13^, reaching undetectable levels in some individuals ^14^. Whether these antibodies are likely to be protective against subsequent infections is becoming clearer. Anti-SARS-CoV-2 Spike antibodies generated following exposure to the original viral variant (D614G) exhibit reduced neutralisation potency against the alpha and beta variants of SARS-CoV-2 ^15-17^. Conversely, antibodies generated following infection with the beta variant are able to potently neutralise the original variant^18-20^.

Though there were initial plans made by the Malawi government in April 2020 to implement a national lockdown of social and commercial activities, these were never implemented ^21^. As such, unlike other countries in the region, including Kenya ^22^, South Africa ^23^ and Zimbabwe ^24^, there were no systematic lockdowns and limited curfew restrictions. Schools were officially closed (from 23^rd^ March 2020 to 7^th^ September 2020) and many social gathering settings (including restaurants and places of worship) voluntarily closed or had significantly reduced services ^25,26^. Malawi closed its borders and airports from April 2020 to September 2020, with limited essential traffic allowed in and out of the country ^27^. As of 12^th^ August 2021, 56,952 people were confirmed to have been infected by SARS-CoV-2 in Malawi, resulting in 1,895 deaths and 169,632 people had been fully vaccinated ^3^.

Due to movement restrictions instituted to curb SARS-CoV-2 transmission, conducting large national population-based seroprevalence studies has been a significant challenge. This has led to the use of blood donor and healthcare worker serosurveys to estimate and monitor the extent of the SARS-CoV-2 pandemic in several countries ^7-9,28^. Here, we report results of a national serosurvey using archived serum samples from blood transfusion donors across Malawi from January 2020 through February 2021, generated using a WHO-recommended anti-SARS-CoV-2 Receptor Binding Domain total antibody assay with high sensitivity and specificity, supported by SARS-CoV-2 pseudovirus neutralisation assays to explore variant specificity.

## Methods

### Study setting and population

The Malawi Blood Transfusion Service (MBTS) maintains an archive of sera from all blood donations received in their Malawi facilities (average of 70,000 annually before the COVID-19 pandemic). All blood donors undergo routine screening through a self-administered questionnaire, one-on-one assessment and a mini-health screening by MBTS staff. Donors meeting routine donor ineligibility criteria including age below 16 or above 65 years, haemoglobin below 12.5g/dl, past medical history suggestive of HIV, hepatitis or syphilis, past or present history of renal, cardiovascular, central nervous system and metabolic disorders, are excluded from donating ^29^. All donated samples are screened for Transfusion Transmissible Infections (TTI; including HIV). Sera are archived at −80°C and retained by MBTS for up to 5 years for retrospective analysis purposes. MBTS blood donation services include both static and mobile sites. For our analysis, we have selected samples from all four defined regions within Malawi: Mzuzu is the capital of the Northern Region and the third largest city, by population, in Malawi (population of 221,272). Lilongwe is Malawi’s capital city located in the Central Region (population of 989,318). Blantyre is the capital of Malawi’s Southern Region (population of 800,264). Balaka is a rural district of 438,379 residents, located in the Eastern Region. All population data are from the 2018 Malawi Population and Housing Census ^30^.

Ethical approval for this study was obtained from the Kamuzu University of Health Sciences Research Ethics Committee (P.09/20/3123). At time of donation, blood donors provide consent to draw blood for transfusion and for use in studies of public health importance. COVID-19 research qualifies as an activity of urgent public health importance.

### Sample selection and processing

Using the MBTS sample archive database, we randomly selected (using STATA’s gsample command) sera collected from HIV-seronegative individuals aged 16-65 years old. Parameters including sex, age, location and time were also extracted from the database for analysis. For Mzuzu, Lilongwe and Blantyre, selected samples were only from static urban donation sites. For Balaka, a largely rural district with smaller total donations, selected samples included donations from both static and mobile sites, including semi-rural and rural.

### Measurement of SARS-CoV-2 antibodies by Enzyme Linked-Immunosorbent Assay

#### SARS-CoV-2 Receptor Binding Domain (RBD) Total Antibody ELISA

We used the WHO-approved WANTAI SARS-CoV-2 Ab commercial Enzyme Linked-Immunosorbent Assay (ELISA) kit to detect total SARS-CoV-2 antibodies, following manufacturer’s instructions (Beijing Wantai Biological Pharmacy Enterprise co., Ltd, China; WS-1096) as reported previously^31^. This ELISA was shown to have the best overall characteristics to detect functional antibodies in different phases and severity of disease ^31^. It is a two-step incubation antigen “sandwich” enzyme immunoassay kit which uses polystyrene microwell strips pre-coated with recombinant SARS-CoV-2 Receptor Binding Domain (RBD) antigen. The optical density (OD) of each well was read at 450nm in a microplate reader (BioTek ELx808, UK) within 10 minutes of adding Stop Solution. Cut-off values were calculated following manufacturer’s instructions. The ratio between a sample’s absorbance and the cut-off were calculated for each sample. Specimens giving a ratio of <0.9 were reported as negative for this assay, a ratio of >1.1 were reported as positive, and a ratio between 0.9-1.1 were reported as borderline. Samples with borderline results were retested using a confirmatory assay described below. The sensitivity and specificity of the assay as independently validated are 97% [95% CI 83.3 to 99.4] and 98% [91.3 to 99.3]), respectively ^32^. The manufacturer’s sensitivity and specificity estimates for the assay are 94% [90.9 to 96.8] and 100% [98.8 to 100], respectively^33^.

#### Confirmatory SARS-CoV-2 Spike 2 and Nucleoprotein IgG antibody ELISA

The COVID-19 IgG RUO commercial ELISA kit (Omega diagnostics, UK) kit uses 96-well microtiter plates pre-coated with purified SARS-COV-2 Spike (S2) and Nucleoprotein (N) antigens to detect SARS-COV-2 IgG. This was performed following manufacturer’s instructions as reported previously^7^. Results were interpreted as described above for the ELISA assay, with a ratio of <0.9 reported as negative, a ratio of >1.1 reported as positive, and a ratio between 0.9-1.1 reported as borderline.

### SARS-CoV-2 Pseudovirus Neutralisation Assay

Samples were pre-screened using an in-house (NICD) spike ELISA^16^, and only samples positive for binding antibodies were screened for neutralization. SARS-CoV-2-pseudotyped lentiviruses were prepared by co-transfecting the HEK 293T cell line with either the SARS-CoV-2 original spike (D614G) or the SARS-CoV-2 Beta spike (L18F, D80A, D215G, K417N, E484K, N501Y, D614G, A701V, 242-244 del) plasmids in conjunction with a firefly luciferase encoding pNL4 lentivirus backbone plasmid. The parental plasmids were kindly provided by Drs Elise Landais and Devin Sok (The International AIDS Vaccine Initiative (IAVI), USA). For the neutralization assay, heat-inactivated seropositive serum samples from blood donors were incubated with the SARS-CoV-2 pseudotyped virus for 1 hour at 37°C, 5% CO_2_.

Subsequently, 1×10^4^ HEK 293T cells engineered to over-express ACE-2, kindly provided by Dr Michael Farzan (Scripps Research), were added and incubated at 37°C, 5% CO_2_ for 72 hours upon which the luminescence of the luciferase gene was measured.

### Statistical analysis

We performed statistical analyses and graphical presentation using R statistical package, version 4.1.0 and GraphPad Prism v9.1.0 (GraphPad Software, LLC). The seroprevalence of SARS-CoV-2 antibodies was adjusted for the independently reported assay sensitivity (97% [95% CI 83.3 to 99.4]) and specificity (98% [95% CI 91.3 to 99.3]) using the bootComb (v1.0.1) R package ^34^. Generalised additive models were used to estimate the seroprevalence curves from Figures 1 and 2. To investigate demographic factors associated with SARS-CoV-2 antibody positivity, a multivariable logistic regression model was developed with sex, age group, location and time as predictors. To obtain smooth regression curves, B-splines ^35^ were used to model the effect of time given the non-linear trend in seroprevalence over time. Effects were considered statistically significant when the *p*-value was less than 0.05.

**Figure 1.**
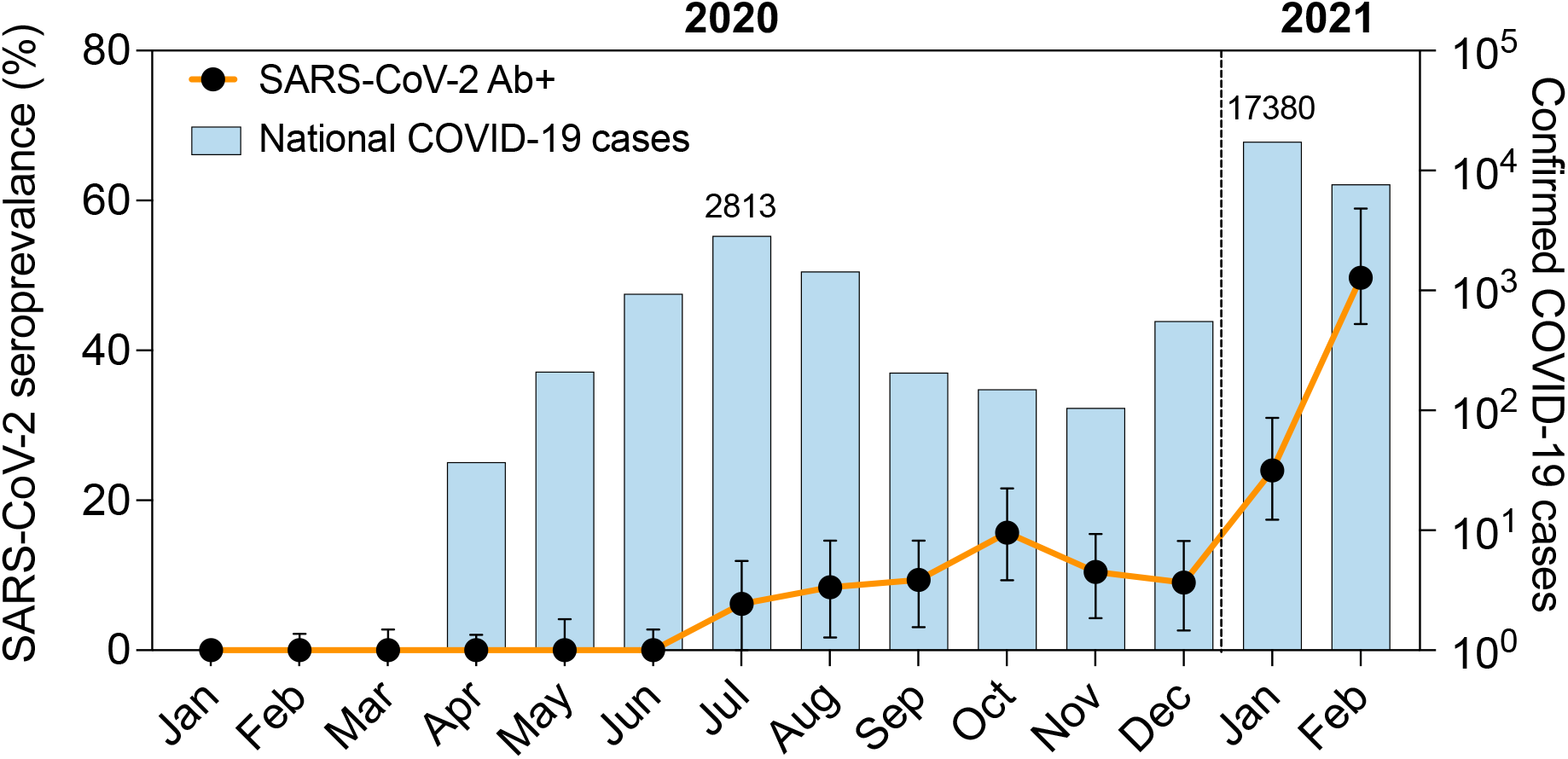
Overall SARS-CoV-2 adjusted seroprevalence over time superimposed on Malawi national PCR-confirmed COVID-19 cases. Black dots (together with 95% CI) are estimated seroprevalence at each timepoint (month), adjusted for assay sensitivity and specificity. The histograms represent confirmed national COVID-19 cases per month, including asymptomatic and symptomatic cases. Case frequency reported above histograms (June 2020 and February 2021) indicates reported cases at the peak of 1st and 2nd wave, respectively. Vertical dotted line defines transition from 2020 to 2021. SARS-CoV-2 = Severe acute respiratory syndrome coronavirus 2, COVID-19 = Coronavirus disease 2019, Ab+ = Positive for detection of anti-SARS-CoV-2 receptor binding domain (RBD) antibody.

**Figure 2.**
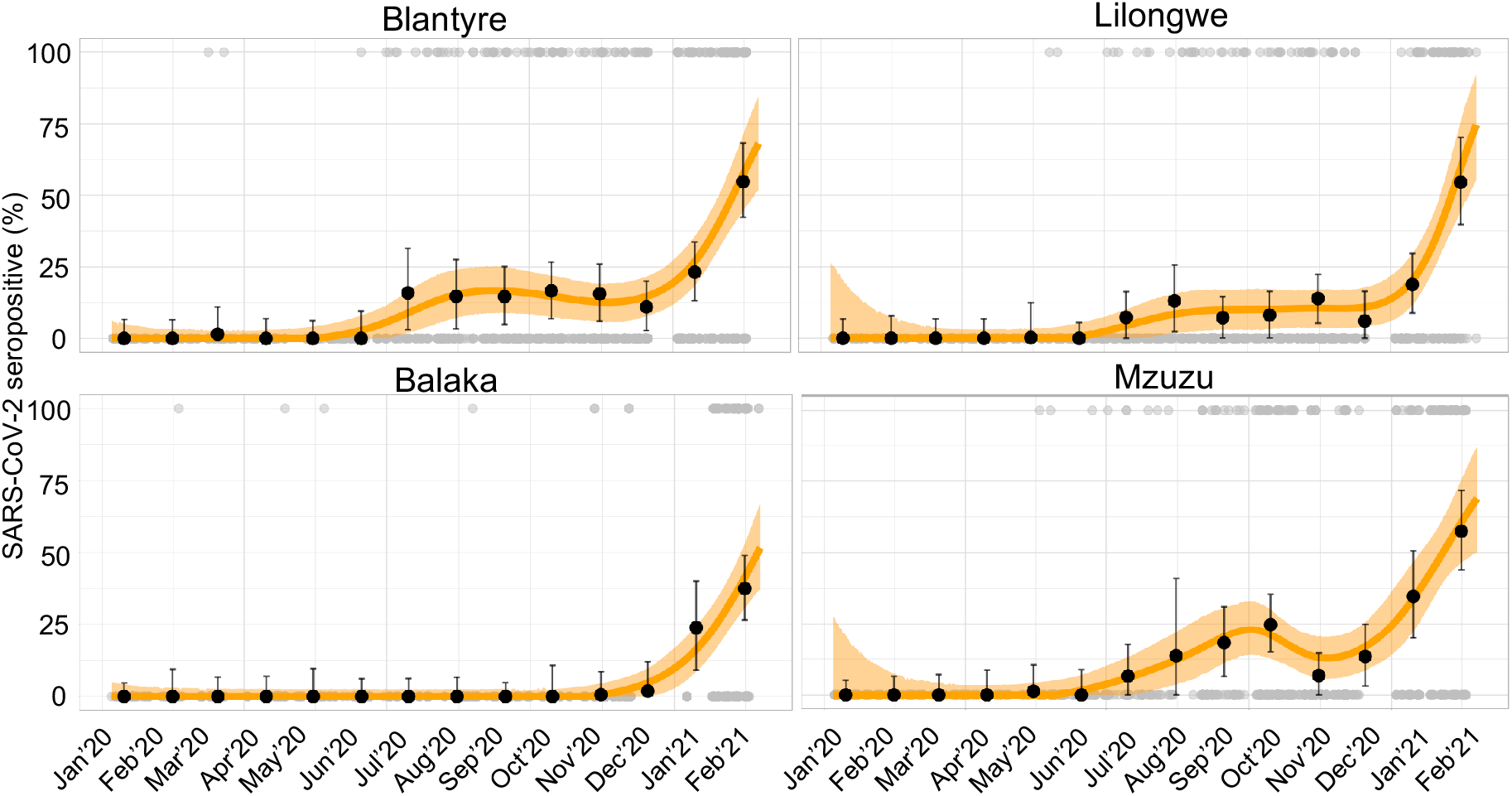
Flexible seroprevalence models. The model fits provide a smoothed estimate of the seroprevalence over time (January 2020 to February 2021) for each location: Blantyre, Lilongwe, Balaka, and Mzuzu. The orange line is the line of best fit for the empirical data, using a smooth generalised additive model, along with light-orange shading indicating 95% CI. Black dots (together with 95% CI) are estimated seroprevalence at each timepoint (month), adjusted for assay sensitivity and specificity. Grey dots (a top and bottom of figures) show the individual-level data, which are either positive (1) or negative (0) for detection of anti- SARS-CoV-2 receptor binding domain (RBD) antibody.

## Results

### Participant demographics

A total of 3,586 blood donor serum samples were selected from the four regional blood transfusion centres. Samples had been collected between January 2020 through February 2021. These included 823 from Mzuzu (Northern region), 946 from Lilongwe (Central region), 830 from Balaka (Eastern region) and 987 from Blantyre (Southern region) (**Table 1**). Compared with 2018 Malawi Population and Housing Census ^30^, by the nature of the demographic of blood donors in Malawi, participants were more commonly male (75% in our study versus 48.5% in the census) and had more persons aged 20 to 49 years (87.3% versus 35.4%).

**Table 1.**
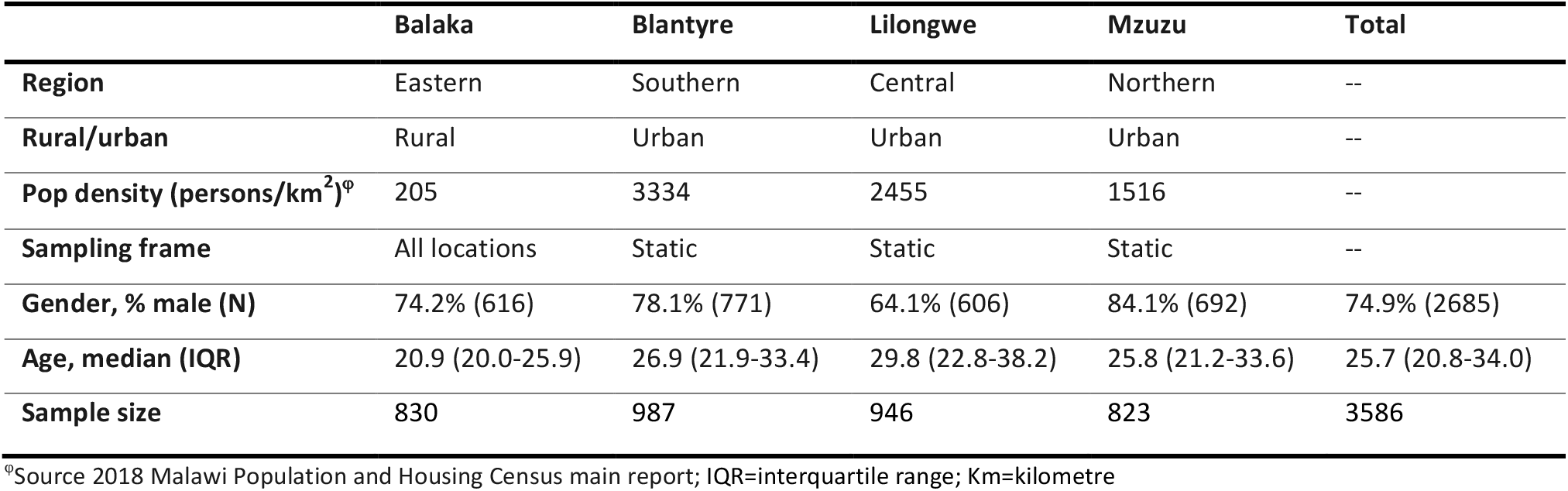
Participant Demographics and site characteristics.

|  | Balaka | Blantyre | Lilongwe | Mzuzu | Total |
| --- | --- | --- | --- | --- | --- |
| **Region** | Eastern | Southern | Central | Northern | -- |
| **Rural/urban** | Rural | Urban | Urban | Urban | -- |
| **Pop density (persons/km<sup>2</sup>)<sup>‡</sup>** | 205 | 3334 | 2455 | 1516 | -- |
| **Sampling frame** | All locations | Static | Static | Static | -- |
| **Gender, % male (N)** | 74.2% (616) | 78.1% (771) | 64.1% (606) | 84.1% (692) | 74.9% (2685) |
| **Age, median (IQR)** | 20.9 (20.0-25.9) | 26.9 (21.9-33.4) | 29.8 (22.8-38.2) | 25.8 (21.2-33.6) | 25.7 (20.8-34.0) |
| **Sample size** | 830 | 987 | 946 | 823 | 3586 |

<sup>‡</sup>Source 2018 Malawi Population and Housing Census main report; IQR=interquartile range; Km=kilometre

### Overall SARS-CoV-2 antibody seroprevalence

Of the 3,586 samples, 456 were positive for anti-SARS-CoV-2 RBD total antibody and 17 were borderline. After re-running the borderline samples using the separate commercial confirmatory ELISA, 13 of the 17 were positive for anti-SARS-CoV-2 S2 and N IgG antibody, resulting in a total 469 anti-SARS-CoV-2 antibody positive samples (crude seroprevalence of 13.1% [95% CI 12.0% to 14.2%]).

As part of quality control procedures, we randomly selected 76 samples that were positive on the primary SARS-CoV-2 ELISA and retested them using the second ELISA targeting S2 and NP IgG. Seventy-one (71) of the 76 samples were positive on the second ELISA, representing a concordance of 93.4% [95% CI 85.3 to 97.8]. We also used a recently published ELISA from the National Institute for Communicable Diseases (NICD, South Africa)^16^ that targets binding antibodies against RBD of the original SARS-CoV-2 variant on a subset of 36 serum samples selected from the seroprevalence peaks. The concordance between the Wantai and NICD assays was 100%.

The overall (all months aggregated) seroprevalence adjusted for the reported independent sensitivity and specificity estimates was 11.2% [95% CI from 5.8% to 14.9%]. In a multivariable logistic regression model, seroprevalence did not vary significantly by sex (*p* = 0.532) or age (*p* > 0.10 for all age groups using those 10-19 years old as reference) but did vary geographically, from 5.5% in Balaka (reference) to 14.6% in Blantyre (*p* < 0.001), 9.8% in Lilongwe (*p* < 0.001) and 14.6% in Mzuzu (*p* < 0.001) (**Table 2**).

**Table 2.** SARS-CoV-2 anti-RBD total antibody seroprevalence in blood donors by participant characteristics and location. This a multivariable logistic regression model with sex, age group, location and time as predictors. B-splines were used to model the effect of time given the non-linear trend in seroprevalence over time.

| Variable | Category | Sample size | Seropositive (frequency) | Unadjusted Seroprevalence (%) | Adjusted Seroprevalence (%) | Estimate | SE | z value | p value |
| --- | --- | --- | --- | --- | --- | --- | --- | --- | --- |
| **Intercept** |  |  |  |  |  | -14.731 | 2.213 | -6.658 | P<0.001 |
| **Gender** | Female | 901 | 108 | 12.0 | 9.5 | Ref | Ref | Ref | Ref |
|  | Male | 2685 | 361 | 13.4 | 11.0 | -0.083 | 0.133 | -0.625 | 0.532 |
| **Age** | 10 - 19<sup>§</sup> | 420 | 35 | 8.3 | 5.6 | Ref | Ref | Ref | Ref |
|  | 20 - 29 | 1844 | 246 | 13.3 | 10.8 | 0.198 | 0.181 | 1.099 | 0.272 |
|  | 30 - 39 | 798 | 112 | 14.0 | 11.6 | 0.215 | 0.201 | 1.067 | 0.286 |
|  | 40 - 49 | 487 | 72 | 14.8 | 12.4 | 0.322 | 0.216 | 1.490 | 0.136 |
|  | 50+ | 37 | 4 | 10.8 | 8.2 | -0.577 | 1.050 | -0.549 | 0.583 |
| **Location** | Balaka | 830 | 64 | 7.7 | 5.5 | Ref | Ref | Ref | Ref |
|  | Blantyre | 987 | 160 | 16.2 | 14.6 | 1.005 | 0.177 | 5.684 | <0.001* |
|  | Lilongwe | 946 | 111 | 11.7 | 9.8 | 0.704 | 0.187 | 3.765 | <0.001* |
|  | Mzuzu | 823 | 134 | 16.3 | 14.6 | 1.187 | 0.183 | 6.496 | <0.001* |

<sup>§</sup> Recruitment limited to blood donors ≥16 years of age
\* Significant based on p-value < 0.05

### Temporal trend of SARS-CoV-2 seroprevalence

The first PCR-confirmed case of COVID-19 identified by Malawi’s national surveillance system was on 2^nd^ April 2021, with the first peak of the national COVID-19 cases being July 2020 (2,813 cases) and subsequently a second peak in January 2021 (17,380 cases) (**Figure 1**). In this blood donor SARS-CoV-2 serosurvey, the first seropositive samples were observed in February 2020 in Balaka (1 sample) and March 2020 in Blantyre (2 samples) (**Figure 2, Table 3 and Supplementary Table 1**). Both unadjusted and adjusted seroprevalence estimates increased with time, with two distinct waves that followed the same temporal trend as the reported national COVID-19 cases (**Figure 1 and Supplementary Table 1**). When aggregating serum samples from all locations, the first wave among donors was observed from July to November 2020 with the peak seroprevalence in October 2020 (15.7% adjusted seroprevalence), while the second wave was observed between December 2020 to February 2021 with peak seroprevalence in February 2021 (49.4%) (**Figure 1, Table 3 and Supplementary Table 1**). However, there were differences in the temporal trend in seroprevalence according to location (**Figure 2, Table 3 and Supplementary Table 1**). Balaka had a 0% adjusted seroprevalence from January to November 2020, subsequently increasing from 1.9% to 37.5% between December 2020 and February 2021. The peak adjusted seroprevalence in the first wave was July 2020 in the Blantyre (15.9%), August 2020 in Lilongwe (13.1%) and October 2020 in Mzuzu (24.8%). Seroprevalence bubble plots at three periods in time demonstrated widespread exposure amongst blood donors across the country in February 2021 compared to the earlier snapshots (**Figure 3**).

**Table 3.** Adjusted seroprevalence, stratified by month and site.

| Yr-month | Balaka % (95% CI) | Blantyre % (95% CI) | Lilongwe % (95% CI) | Mzuzu % (95% CI) | Total % (95% CI) |
| --- | --- | --- | --- | --- | --- |
| 2020-01 | 0.0 (0.0, 4.7) | 0.0 (0.0, 6.6) | 0.0 (0.0, 6.7) | 0.0 (0.0, 5.2) | 0.0 (0.0, 2.4) |
| 2020-02 | 0.0 (0.0, 9.5) | 0.0 (0.0, 6.6) | 0.0 (0.0, 7.9) | 0.0 (0.0, 6.4) | 0.0 (0.0, 2.8) |
| 2020-03 | 0.0 (0.0, 6.8) | 1.5 (0.0, 11.1) | 0.0 (0.0, 6.7) | 0.0 (0.0, 7.1) | 0.0 (0.0, 2.8) |
| 2020-04 | 0.0 (0.0, 7.0) | 0.0 (0.0, 7.1) | 0.0 (0.0, 6.8) | 0.0 (0.0, 8.6) | 0.0 (0.0, 2.8) |
| 2020-05 | 0.0 (0.0, 9.5) | 0.0 (0.0, 6.1) | 0.2 (0.0, 12.5) | 1.3 (0.0, 10.4) | 0.0 (0.0, 3.9) |
| 2020-06 | 0.0 (0.0, 6.2) | 0.0 (0.0, 9.6) | 0.0 (0.0, 5.4) | 0.0 (0.0, 8.8) | 0.0 (0.0, 2.7) |
| 2020-07 | 0.0 (0.0, 6.2) | 15.9 (2.9, 31.2) | 7.3 (0.0, 16.4) | 6.6 (0.0, 17.6) | 6.2 (0.5, 11.4) |
| 2020-08 | 0.0 (0.0, 6.7) | 14.7 (3.3, 27.6) | 13.1 (2.2, 25.6) | 13.7 (0.0, 40.9) | 8.4 (1.7, 14.4) |
| 2020-09 | 0.0 (0.0, 4.7) | 14.6 (4.8, 25.0) | 7.2 (0.1, 14.5) | 18.2 (6.6, 31.3) | 9.4 (3.0, 14.9) |
| 2020-10 | 0.0 (0.0, 10.8) | 16.7 (7.0, 26.8) | 8.1 (0.2, 16.5) | 24.8 (14.8, 35.2) | 15.7 (8.9, 22.0) |
| 2020-11 | 0.6 (0.0, 8.7) | 15.6 (6.0, 25.9) | 13.9 (5.4, 22.6) | 6.8 (0.0, 14.7) | 10.4 (4.0, 15.8) |
| 2020-12 | 1.9 (0.0, 12.0) | 11.0 (2.5, 19.8) | 6.0 (0.0, 16.5) | 13.4 (3.1, 25.0) | 9.1 (2.7, 14.9) |
| 2021-01 | 23.9 (9.4, 40.2) | 23.2 (13.2, 33.7) | 18.8 (8.8, 29.7) | 34.7 (19.9, 50.7) | 24.0 (16.7, 31.2) |
| 2021-02 | 37.5 (26.7, 49.0) | 54.8 (42.4, 67.5) | 54.5 (39.8, 70.4) | 57.5 (44.4, 72.1) | 49.7 (42.3, 58.8) |

**Figure 3.**
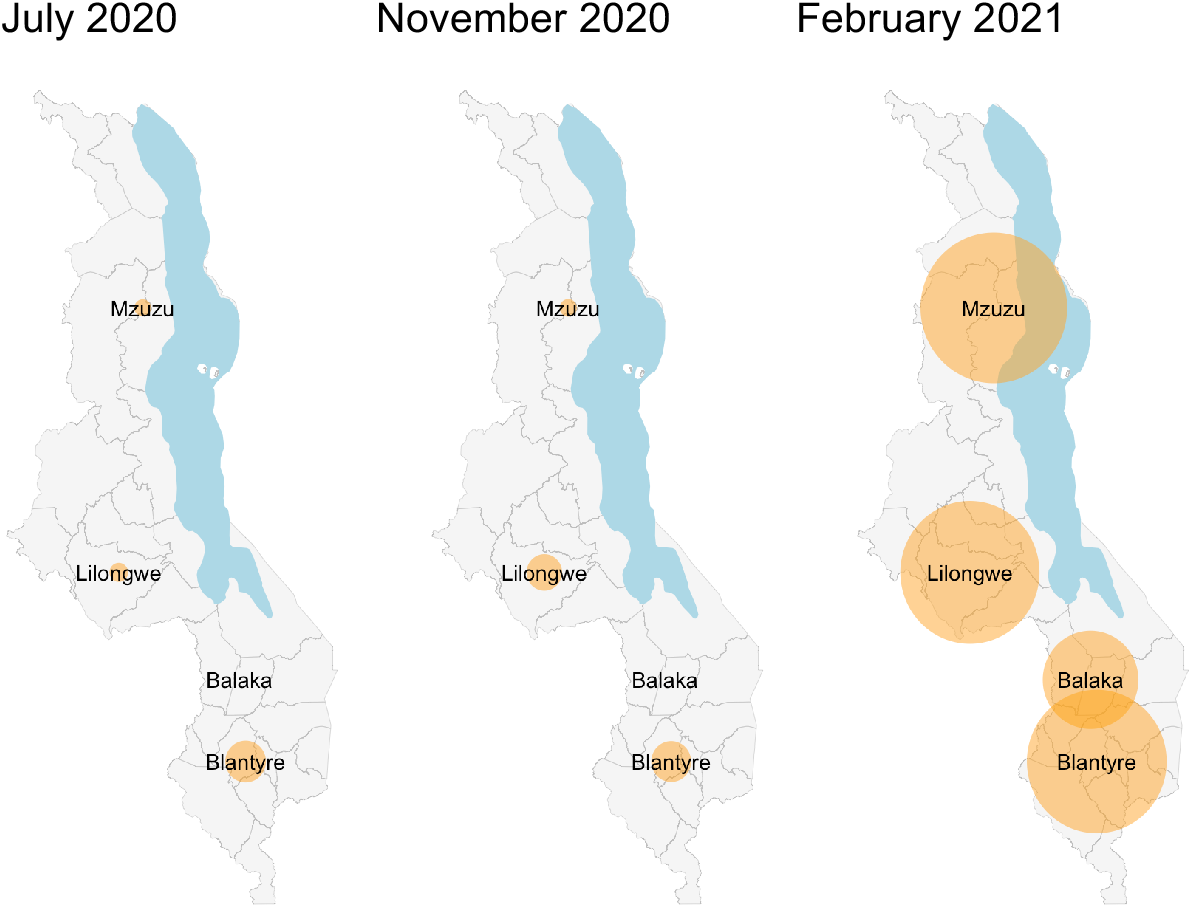
Seroprevalence bubble plot for overall seroprevalence at three snapshots in time. The size of the bubble is proportional to the prevalence of samples positive for anti-SARS-CoV-2 receptor binding domain (RBD) antibody within each location.

### Estimating predominant SARS-CoV-2 variants driving seroprevalence

Limited evidence suggests that Malawi’s second wave was driven by the Beta variant, with data deposited on GISAID showing that the variant was identified in 96% (152/159) of the samples sequenced from Malawi from January 2021 ^36^. As it has been shown that antibodies elicited by the original variant (D614G WT) are less potent against the Beta variant^16,19^, we reasoned that differential neutralisation potency by SARS-CoV-2 antibodies could be used to estimate the predominance of specific variants driving seroprevalence. We therefore measured neutralisation antibody responses against D614G WT and Beta variant in randomly selected seropositive sera collected at the peak of the first wave (Wave 1) in June to October 2020 compared to those collected in February 2021 (Wave 2). Wave 1 sera was more potent against D614G WT than the Beta variant, while Wave 2 sera was relatively more potent against the Beta variant than D614G WT (**Figure 4a-b**). Only 22% (2/9) of the Wave 1 sera was more potent against the Beta variant than D614G WT, while 47% 8/17 of the Wave 2 sera was more potent against the Beta variant than D614G WT (**Figure 4b-c**). In contrast, 78% (7/9) of the Wave 1 sera was more potent against D614G WT than the Beta variant, while 18% 3/17 of the Wave 2 sera was more potent against D614G WT than the Beta variant (**Figure 4c**). Furthermore, 35% (6/17) of the Wave 2 sera showed no neutralisation against the Beta variant or D614G WT (**Figure 4c**). Collectively, these results support existing sequencing evidence that Malawi’s second SARS-CoV-2 epidemic wave was driven by the Beta variant.

**Figure 4.**
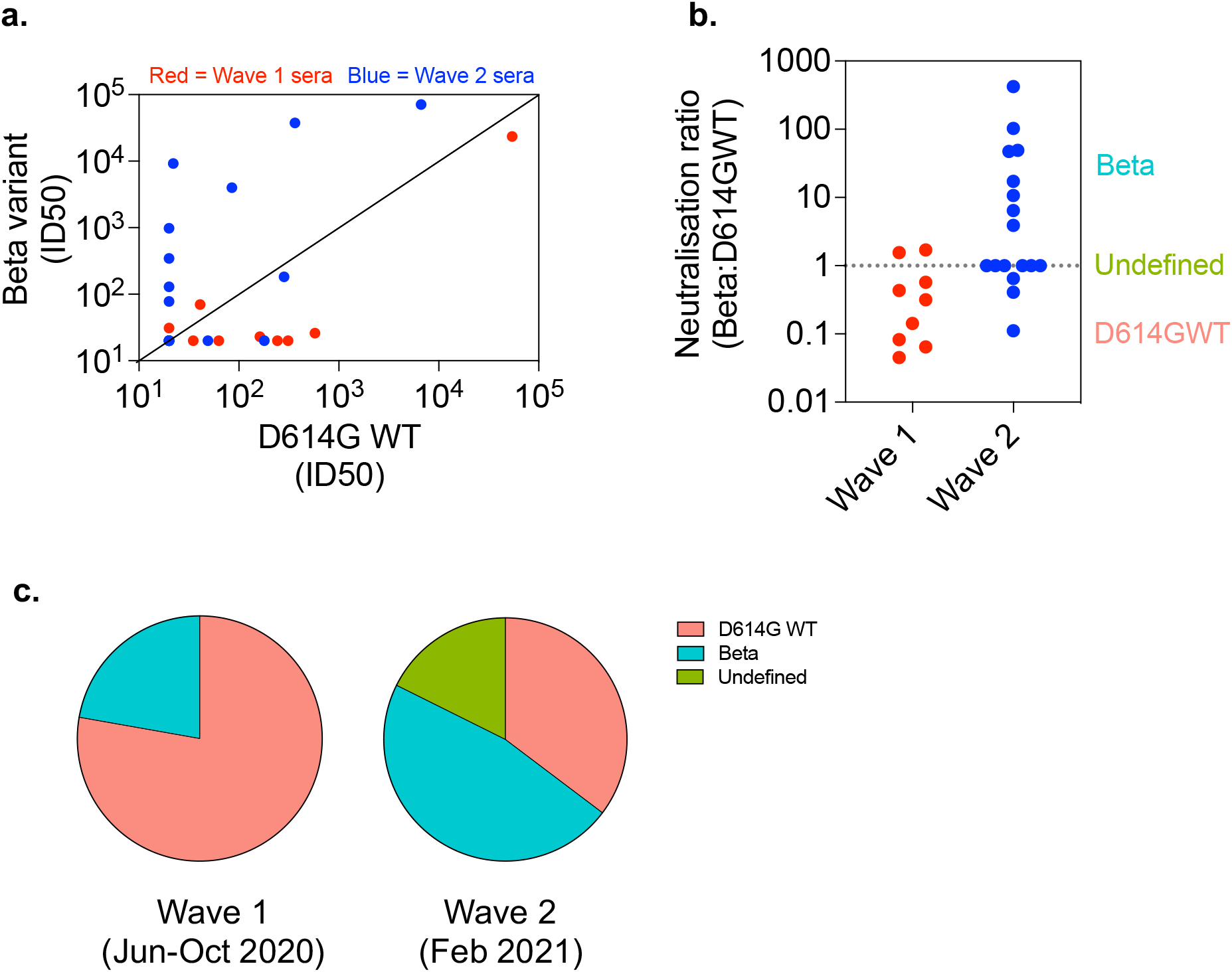
SARS-CoV-2 neutralisation of D614G WT and Beta variant using sera from the first and second epidemic waves. Anti-RBD seropositive sera from the SARS-CoV-2 epidemic Wave 1 and 2 were subjected to a SARS-CoV-2 pseudovirus neutralisation assay. a) Correlation of SARS-CoV-2 antibody neutralisation potency against D614G WT and Beta variant. b) Magnitude of neutralisation potency of Wave 1 (n=9) and Wave 2 (n=17) sera against D614G WT and Beta variant. Sera with a ratio of >1 are classified as Beta variant-induced antibodies, <1 as D614G WT-induced antibodies and 1 as undefined. c) Proportion of Wave 1 and Wave 2 sera with relatively higher neutralisation potency against D614G WT and Beta variant. The threshold of detection for the neutralization assay is ID_50_>20. RBD, Receptor Binding Domain.

## Discussion

Our study provides important insights into the dynamics of the COVID-19 epidemic in both urban and rural Malawi, with likely relevance to the southern Africa region. The seroprevalence estimates from this blood donor serosurvey are very high compared to the reported national surveillance figures for confirmed asymptomatic and symptomatic cases, but they do reflect the emergence and magnitude of the first and second COVID-19 epidemic waves in Malawi. They also provide evidence against the speculation that SARS-CoV-2 had been circulating more widely in Malawi before the first detected case in April 2020 or that circulation of endemic coronaviruses ^10,11^ had generated cross-reactive antibody responses. Data from pseudovirus neutralisation assays suggests that the second wave was predominantly driven by the Beta variant, as sera from the first wave had poor neutralising activity against the Beta variant of SARS-CoV-2. Collectively, our findings provide further support to the emerging data that in terms of hospitalised disease and death, the COVID-19 pandemic has so far been attenuated in sub-Saharan Africa, despite high population exposure to SARS-CoV-2.

We report a high seroprevalence of 49.7% nationally in February 2021, mirroring the upsurge in confirmed COVID-19 cases, hospitalisations and death in Malawi experienced during this period ^3,6^. This is consistent with a study in South African blood donors that reported seroprevalence rates ranging from 31.8% to 62.5% in January 2021 ^37^. So far, the epidemic trajectory in Malawi as determined by the national confirmed COVID-19 cases, deaths and hospitalisations has paralleled the epidemic trajectory in South Africa ^3^. This is likely due to shared borders and the resulting frequent human migration between the two countries, specifically in the last 12 months during which there has been an influx of groups of returning residents from South Africa to Malawi. This movement of people coincided with rapid surges in COVID-19 cases in the first and second epidemic waves in Malawi ^38^. These findings support the need for a consolidated and standardised regional public health effort in effectively addressing the COVID-19 pandemic in southern Africa to reduce the risk of further epidemic waves across the region.

Genomic surveillance data from Malawi and South Africa suggests that this high seroprevalence in the second wave is, at least in part, driven by the emergence of the Beta variant ^36^. In agreement, our neutralisation data showed that sera collected from the second wave (February 2021) was better at neutralising the Beta variant than that from the first wave (June to October 2020), indicating antibody responses driven by infection with the Beta variant rather than the original variant (D614G WT). This is consistent with previous studies showing that homologous convalescent sera retain higher *in vitro* neutralisation potency than heterogous convalescent sera against the infecting virus ^15-20^. It is noteworthy that 35% of the Wave two sera that were positive for antibodies to the SARS-CoV-2 RBD showed no neutralisation against the Beta variant or D614G WT, highlighting a disconnect between qualitative antibody detection and functional activity. This discordance could be due to low anti-RBD antibody titers as they have been shown to be associated with poor *in vitro* neutratisalion ^19,20^.

The temporal kinetics of the SARS-CoV-2 seroprevalence in this blood donor serosurvey suggests that the first epidemic wave was largely confined to urban areas. This differs to the second wave, which was not only more rapid in its growth but also in its geographic spread, as shown by the high seroprevalence even in the rural areas. This is consistent with national surveillance reports showing that confirmed COVID-19 cases in the first wave were primarily from the three major cities, Blantyre, Lilongwe and Mzuzu, but in the second wave there was an increase in reported cases in rural areas ^39,40^. Also noteworthy is the earlier seroprevalence peak in Blantyre and Lilongwe, compared to the northern city of Mzuzu. Blantyre and Lilongwe are the largest commercial centres in Malawi with multiple connection to South Africa through international airports and bus stations. Mzuzu is the hub for inter-country trade and migration between Tanzania and Malawi and it is plausible that some of the late epidemic surge in Mzuzu was due to cross border movement of persons between Tanzania and Malawi.

This study has considerable strengths, including consistent monthly sampling, use of well-validated assays and national geographical representation. There are however some important limitations. First, inherent in using a blood donor sampling frame is that the convenience sampling was not representative of the general population, skewed towards males and individuals aged 18 to 45 years old. In addition, we were unable to evaluate the association between the behaviour of blood donors and risk of acquisition to SARS-CoV-2 infection, as this may bias the measured results away from the true population seropositivity. Though random sampling from the larger population would have been ideal, restrictions on movement and gathering make these study designs extremely challenging to implement. Second, in some individuals, SARS-CoV-2 antibodies wane over time to undetectable levels leading to false negatives, especially in those who had asymptomatic COVID-19 ^12-14^. Therefore, the seroprevalence estimates are likely an underestimate of the cumulative exposure within this sampled population. However, the selection bias and waning of antibody levels is unlikely to substantially alter the temporal trends reported in this study.

In conclusion, we report a dramatic rise in SARS-CoV-2 seroprevalence from 6.2% in July 2020 to 49.7% in February 2021 in healthy blood donors as Malawi experienced the first and second COVID-19 epidemic waves, likely driven initially by the original variant (D614G WT) and then the Beta variant. Based on data showing prior exposure to SARS-CoV-2 augments COVID-19 vaccine responses ^41-44^, prioritisation of the first COVID-19 vaccine dose in settings with high seroprevalence could help maximise the benefit of the limited available COVID-19 vaccine doses. The dynamics of SARS-CoV-2 exposure will therefore need to be taken into account in the formulation of COVID-19 vaccination policy in Malawi and across the region.

## Data Availability

All data is available upon request

## Acknowledgements

The authors thank all blood donors whose samples are used in this study, the staff of the Malawi-Liverpool-Wellcome Trust Clinical Research Programme (MLW) and Malawi Blood Transfusion Services (MBTS) for their support and co-operation during the study.

## Author Contributions

KCJ, TDS, RSH, NF, BM, TM, AM and NK designed the study. KCJ, TDS, TM, GK, AL, NM and PLM supervised the work. JM, CB, AS, FA, ZM, TM, NM, AS and TMG processed all samples and carried out all laboratory-based assays. MYRH, BM and JM carried out data management and statistical analysis. KCJ, TDS, JM, RSH and NF wrote the initial manuscript draft. All authors reviewed and approved the final manuscript.

